# Functional analysis of NANOS protein variants identified in men with infertility employing zebrafish embryos

**DOI:** 10.64898/2026.10.20.26366003

**Authors:** Shantanu Madiwale, Antra Gupta, Leon Schmidthuis, Csilla Krausz, Viola Bonini, Sabine Kliesch, Corinna Friedrich, Frank Tüttelmann, Erez Raz

**Affiliations:** Institute of Cell Biology, ZMBE, Von-Esmarch-Straße 56, 48149 Muenster, Germany; Department of Experimental and Clinical Biomedical Sciences “Mario Serio”, University of Florence, 50139 Florence, Italy; Department of Andrology, Fundacio Puigvert, Instituto de Investigaciones Biomedicas Sant Pau (IIB-Sant Pau) 08025 Barcelona, Catalonia, Spain; Department of Andrology, Centre of Reproductive Medicine and Andrology, University Hospital Münster, 48149 Münster, Germany; Institute of Reproductive Genetics (IRG), Centre of Medical Genetics (CMG), University of Münster, 48149 Münster, Germany; Max-Planck-Institute for Molecular Biomedicine, Roentgenstraße 20, 48149 Muenster, Germany

**Keywords:** NANOS, azoospermia, male infertility, non-obstructive azoospermia, exome sequencing, zebrafish, Functional variant assay

## Abstract

**Study question:** Are vertebrate NANOS proteins associated with human fertility, and can a rapid *in vivo* assay in zebrafish be established to evaluate the biological activity of human *NANOS* variants?

**Summary answer:** Combining a rapid *in vivo* assay we developed for Nanos protein activity and patient-derived genetic data, we identified *NANOS* variants that could contribute to male infertility.

**What is known already:** Male infertility affects approximately 17% of the population, but linking specific gene variants to this condition remains challenging. NANOS proteins are essential regulators of germ cell development in different model organisms; however, a rapid and robust *in vivo* assay for evaluating their activity in the context of human male infertility is currently lacking.

**Study design, size, duration:** Exome/genome data from 3,338 men included in the Male Reproductive Genomics (MERGE) cohort were examined in this study. Of these, 2038 men showed severely impaired spermatogenesis but were otherwise healthy. Ninety-Six men with intact spermatogenesis were included in the study as controls. Additionally, the study included a targeted analysis of *NANOS* variants in 44 patients from the IMIGC cohort.

**Participants/materials, setting, methods:** The exome/genome data were screened for rare, stop-gain, frameshift, splice site, as well as missense variants in *NANOS1*, *NANOS2*, and *NANOS3*. Human amino acid substitutions were introduced at the corresponding positions in the zebrafish proteins. Global co-expression of wild-type Dnd1 and Nanos3 in zebrafish embryos induces a germ cell-like fate in somatic cells, resulting in severe developmental defects and embryonic lethality. Based on this principle, we established a rapid *in vivo* assay in which wild-type Dnd1 was co-expressed with wild-type or variant Nanos2 and Nanos3 proteins in live zebrafish embryos, expecting early embryonic death if a variant protein is functional. This assay was used to evaluate the activity of *NANOS2* and *NANOS3* variants identified in men with infertility.

**Main results and the role of chance:** In human exome/genome sequencing data, we identified one heterozygous missense variant in NANOS2 and two heterozygous missense variants in NANOS3. The function of the variants was examined in the zebrafish using the *in vivo* assay we developed. Co-expression of wild-type Dnd1 with selected Nanos2 or Nanos3 variants identified in infertile men resulted in defects in induction of the germ cell-like phenotype, demonstrating the power of the biological readout for evaluating the potential effects of patient-derived variants on NANOS protein activity. Together, our analysis enabled the evaluation of single nucleotide variants, whose impact on protein function is difficult to predict, and allowed us to distinguish variants with little or no effect on protein function from those that strongly reduce activity and may potentially contribute to infertility.

**Limitations, reasons for caution:** The assay requires access to zebrafish embryos and basic imaging equipment. Although the functional conservation of NANOS proteins between zebrafish and humans is supported by previous studies, species-specific differences may influence protein activity. Therefore, the zebrafish assay should be considered one component of a broader framework for evaluating *NANOS* variants as contributing factor for infertility.

**Wider implications of the findings:** Using *NANOS2* and *NANOS3* as examples, we demonstrate that integrating clinical genetic data with functional cell and developmental biology can help establish links between candidate disease genes and human fertility. This strategy may apply to other germline-specific genes whose ectopic expression produces measurable developmental phenotypes in zebrafish embryos.

## Introduction

Male infertility affects approximately 17% of the population, with the complete absence of spermatozoa in the semen (azoospermia) representing the most severe manifestation of the disease (World Health Organization (WHO), 2023). Conventional fertility diagnostic approaches, including detailed investigation of the history of the disease, focused physical examination, and selective laboratory tests, often fail to reveal the actual cause of male infertility cases linked to defects in spermatogenesis (Agarwal et al., 2021; Eisenberg et al., 2023; Tournaye et al., 2017; Tüttelmann et al., 2025). Thus, the underlying cellular and molecular etiologies for infertility frequently remain elusive, although it is believed that the basis for majority of severe cases is genetic (Aitken and Norman, 2026; Krausz and Riera-Escamilla, 2018; Tüttelmann et al., 2018). Indeed, recent advances in large-scale genetic analysis of patient-derived samples have facilitated the identification of multiple gene-specific variants potentially causative for male infertility (Alhathal et al., 2020; Fakhro et al., 2018; Riera-Escamilla et al., 2025; Stallmeyer et al., 2025; Westerich et al., 2023a). The impact of specific gene variants can be evaluated by the use of animal models in a physiologically relevant *in vivo* context. The zebrafish is a particularly well-suited vertebrate model for such functional assays, as germline development is well-characterized and amenable to genetic manipulation and imaging (Weidinger et al., 1999; Raz, 2003; Gross-Thebing and Raz, 2020; Beer and Draper, 2013; Aharon and Marlow, 2021; Marlow, 2015; Abrams and Mullins, 2009; Elkouby and Mullins, 2017).

We have previously shown that the two proteins Dead end1 (Dnd1) and Nanos3 (Nos3) play a critical role in zebrafish germ-cell development (Gross-Thebing et al., 2017; Slanchev et al., 2009; Weidinger et al., 1999; Westerich et al., 2023b). Indeed, the two RNA-binding proteins were found to be essential in different stages of germ cell development in different organisms, from early germline specification to gametogenesis (Horvay et al., 2006; Julaton and Reijo Pera, 2011; Kobayashi et al., 1996; Ruthig et al., 2021; Subramaniam and Seydoux, 1999; Tsuda et al., 2003). In addition to the importance of the proteins in germ cell development in model organisms, we have shown that selected Dnd1 variants identified in men with infertility due to spermatogenic failure exhibit reduced activity when assayed in zebrafish embryos (Westerich et al., 2023a).

In humans, three NANOS paralogs -NANOS1, NANOS2, and NANOS3 -perform distinct functions (Ilaslan et al., 2022; Julaton and Reijo Pera, 2011; Kunik et al., 2025; Kusz et al., 2009). In mice, disruption of Nanos2 causes male sterility (Sada et al., 2009; Saga, 2010; Suzuki et al., 2007a; Tsuda et al., 2003, 2006). Similarly, Nanos2 is proposed to play a crucial role in germline differentiation in medaka and zebrafish (Aoki et al., 2009; Beer and Draper, 2013). Loss of Nanos3 results in the complete absence of germ cells in zebrafish and mice (Köprunner et al., 2001; Lolicato et al., 2008; Tsuda et al., 2003). While absence or disruption of Nanos2 and Nanos3 leads to infertility in mice, Nanos1 is expressed in central nervous system and adult brain and is dispensable for normal development (Haraguchi et al., 2003; Saba et al., 2014; Suzuki et al., 2007a; Tsuda et al., 2003).

Using *in vitro* assays, a homozygous NANOS3 variant p.E120K in the conserved zinc finger domain, impairs germ-cell viability and has been linked to premature ovarian insufficiency (Santos et al., 2014). In addition, E120K and R153W mutations impair NANOS3-mediated repression of RNA translation in cell lines (Sajek et al., 2018). Although *NANOS2* gene variants within the highly conserved RNA-binding domain were identified in men with infertility, their functional relevance has not been established (Kusz et al., 2009). Together, while exome/genome sequencing of men and women with infertility can identify multiple variants, high-throughput *in vivo* assays determining the function of the protein variants can help distinguish pathogenic from benign variants.

In this study, we present a novel assay in which we employed zebrafish embryos as a sensitive *in vivo* model to assist in the functional analysis of *NANOS* gene variants. The assay we developed is based on the finding that expression of germline RNAs in somatic cells results in their transformation into germ cells (Wang et al., 2023) and on the later finding made in Medaka fish, where it was shown that co-expression of Nanos3 and Dnd1 is sufficient for the transformation of somatic cells into PGC-like cells (Nishimura and Fujimoto, 2025). Based on this principle, we established a high-throughput *in vivo* screening method to evaluate potentially causative and non-causative *NANOS2* and *NANOS3* variants found in men with infertility. By employing this method, we were able to identify Nanos protein variants that affect protein function, potentially further linking NANOS function to male fertility in humans.

## Results

### Zebrafish Nanos2 and Nanos3 induce PGC-like fate and suppress somatic differentiation

We have found that as shown in medaka fish, global co-expression of wild-type Nanos3 (Nanos3^WT^) and wild-type Dnd1 (Dnd1^WT^) in zebrafish embryos results in the formation of PGC-like cells and a lack of somatic-cell differentiation. This treatment resulted in pronounced abnormalities in embryonic development and eventual lethality (**Fig. 1 A**), clearly distinguishable from control embryos that developed and hatched normally by 72 hours post fertilization (hpf) (Compare treated and control in **Fig. 1 A**). To assess whether the other Nanos paralogs, i.e. Nanos1 and Nanos2 (**Fig. 1 B**) can induce PGC-like fate when co-expressed with Dnd1^WT^, we conducted an experiment presented in **Fig. 1 A**. Indeed, in this assay, the wild-type Nanos2 (Nanos2^WT^) protein exhibited activity comparable to that of Nanos3^WT^, while Nanos1^WT^ did not (**Fig. 1 C**). A very small fraction of embryos co-injected with *dnd1* and *nanos2* mRNAs successfully hatched; however, these larvae exhibited pronounced morphological abnormalities. (**Supplementary Fig. S1A**).

**Figure 1:**
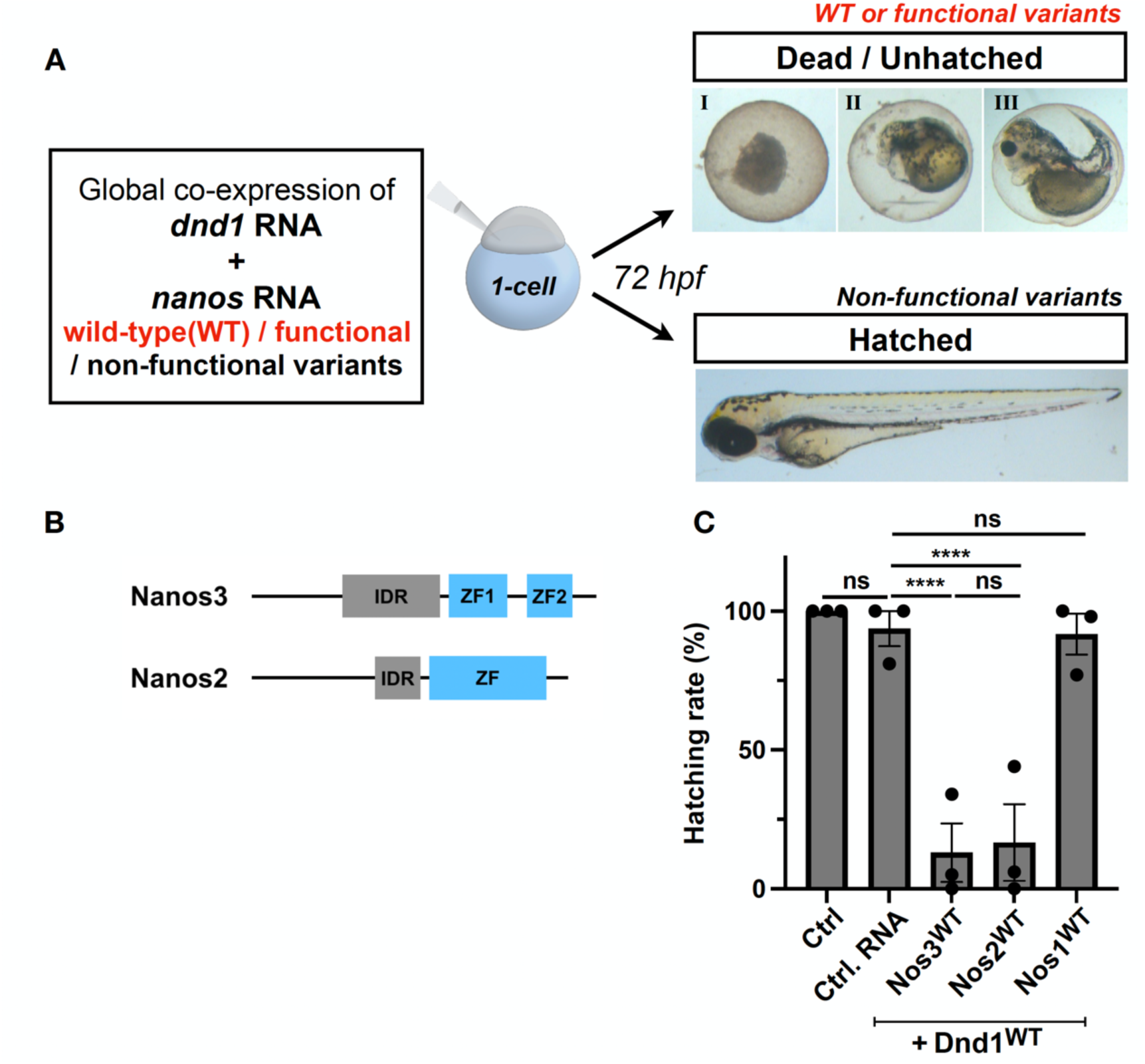
Establishment of a robust and rapid *in vivo* assay for Nanos2 and Nanos3 protein activity. (A) Schematic illustration of the experimental setup. One-cell-stage embryos were co-injected with either *nanos1*, *nanos2*, *nanos3* or *control* mRNA, along with WT *dnd1* mRNA. Imaging of the embryos and evaluation of hatching rate was performed at 72 hours post fertilization (hpf). Representative images of hatched and defective embryos at 72 hpf. For the Nanos3 experiment unhatched embryos were categorized based on morphology - Type I = 91/ 115 embryos, Type II = 13/115, Type III = 11/115. (B) Illustration of different domains of the zebrafish Nanos2 and Nanos3 proteins. IDR – Intrinsically disordered region; ZF – Zinc finger domain. (C) Quantification of hatching rate at 72 hpf for embryos globally co-expressing RNA encoding for the wild-type version of Dnd1 and either of the Nanos proteins or control. (N=3, Control n = 101; Control RNA n = 186; Nos3^WT^ n = 132; Nos2^WT^ n = 117; Nos1^WT^ n = 154). ns : Not significant (P>0.05), * : P ≤ 0.05, ** : P ≤ 0.01, *** : P ≤ 0.001, **** : P ≤ 0.0001.

The functional conservation of Nanos2 and Nanos3 was further demonstrated by the ability of both proteins to reverse the effect of a morpholino antisense oligonucleotide (MO) directed inhibition of Nanos3 function. In this experiment, we co-injected the MO directed against *nanos3* RNA and RNAs lacking the MO-binding sites that encode for Nanos1^WT^, Nanos2^WT^, or Nanos3^WT^ proteins. Consistent with the effect on embryonic development presented above, expression of Nanos2^WT^ and Nanos3^WT^ restored the number of PGCs at the developing gonad region to its normal level, while the Nanos1^WT^ protein was much less efficient in this assay (**Supplementary Fig. S1B**).

Taking these results together, this model system provides a valuable tool to analyze human *NANOS2* and *NANOS3* variants for their protein function.

### Identification of rare *NANOS2* and *NANOS3* variants in men with infertility

In the exome/genome sequencing data from the Male Reproductive Genomics (MERGE) study cohort, we identified two rare heterozygous variants in *NANOS3* (NM_001098622.2) conserved between human and zebrafish. One rare, conserved and heterozygous *NANOS2* variant (NM_001029861) was identified through the International Male Infertility Genomics Consortium (IMIGC) (**Table 1**).

**Table 1:** Genetic and clinical data of men with variants in *NANOS2* and *NANOS*.

| Subject ID | Age (years); country of origin | Semen analysis | Testicular volume (left/right) [ml] | FSH [IU/l] | LH [IU/l] | T [nmol/l] | Variant (cDNA) | Variant (protein) | Variant (zebrafish) | CADD score | MAF gnomAD v4.1.1 |
|---|---|---|---|---|---|---|---|---|---|---|---|
| ***NANOS3*** |  |  |  |  |  |  |  |  |  |  |  |
| M3285 | 25-30; Italy | Severe oligozoospermia | 14/17 | 26.7 | 8.8 | 16 | c.155C>T | p.(Pro52Leu) | Pro71Leu | 5.9 | 0.00002739 |
| M2941 | 51-56; Germany | Azoospermia | 19/40 | 15 | 3.5 | 20.6 | c.253G>A | p.(Glu85Lys) | Glu100Lys | 31 | 0.0000006205 |
| ***NANOS2*** |  |  |  |  |  |  |  |  |  |  |  |
| NOA5 | na | Azoospermia* | 12/10 | 21.8 | 7.5 | 19.7 | c.208G>A | p.(Gly70Arg) | Gly90Arg | 32 | 0.0001413 |

Abbreviations: FSH: follicle stimulating hormone, LH: luteinising hormone, T: testosterone, CADD: Combined Annotation Dependent Depletion, MAF: minor allele frequency, na: not available \* Testistcular sperm extraction (TESE) was performed and no spermatozoa retrieved.
Reference values: testicular volume >12 ml each, FSH: 1–7 IU/l, LH: 2–10 IU/l, T > 12 nmol/l; bold font marks values outside the normal range.

Individual M3285 with the missense variant c.155C>T p.(Pro52Leu) in *NANOS3* had severe oligozoospermia (HP: 0034818, 2 million sperm in the ejaculate). M2941 with the missense variant c.253G>A p.(Glu85Lys) in *NANOS3* displayed azoospermia (HP: 0000027, no spermatozoa in the ejaculate). Both men had elevated follicle stimulating hormone (FSH) levels while luteinizing hormone (LH), testosterone, and testicular volumes were within the reference values.

Individual NOA5 with the missense variant c.208G>A p.(Gly70Arg) in *NANOS2* presented with azoospermia with high FSH and normal LH and testosterone levels and slightly hypotrophic testes (Table 1).

No men with homozygous or two heterozygous variants meeting the selection criteria were identified. An alternative genetic cause of male infertility, such as variants in valid disease genes, was ruled out in the three men.

### Evaluation of the activity of human NANOS3 variants in zebrafish embryos suggests that the NANOS3^E85K^ is a loss-of-function version of the protein

As mentioned above, two NANOS3 variants, p.Pro52Leu (P52L) and p.Glu85Lys (E85K) were identified in men with infertility (**Table 1**). To assess the activity of these proteins, human NANOS3 and zebrafish Nanos3 protein sequences were aligned, allowing the identification of the amino acids in the zebrafish Nanos3 protein that correspond to the altered amino acids in the human patient (**Fig. 2 A**). We therefore generated zebrafish Nanos3 P71L (Nanos3^P71L^) and E100K (Nanos3^E100K^) variants, corresponding to human NANOS3 variant P52L and E85K, respectively (**Fig. 2 A**). Each of the Nanos3 variants was then co-expressed with Dnd1^WT^, and hatching rates at 72 hpf were assessed as outlined in the **Fig. 1 A**. Co-expression of Nanos3^WT^ with Dnd1^WT^ induced strong embryonic malformations and lethality, consistent with robust induction of PGC-like cells and disruption of normal development (**Fig. 2 B**). Notably, the Nanos3^P71L^ variant caused embryonic death with similar efficiency to Nanos3^WT^ (**Fig. 2 B**), suggesting that the function of this form of the protein (the NANOS3^P52L^ human variant) is intact and is thus less likely to contribute to the patient’s infertility. The preservation of protein function in this case is not unexpected, as the Combined Annotation Dependent Depletion (CADD) score is low (5.9) and as the residue at position 71 lies within the non-conserved intrinsically disordered region (IDR) of Nanos3 (**Table 1**, **Fig. 2 A**) (Zarin et al., 2019).

**Figure 2:**
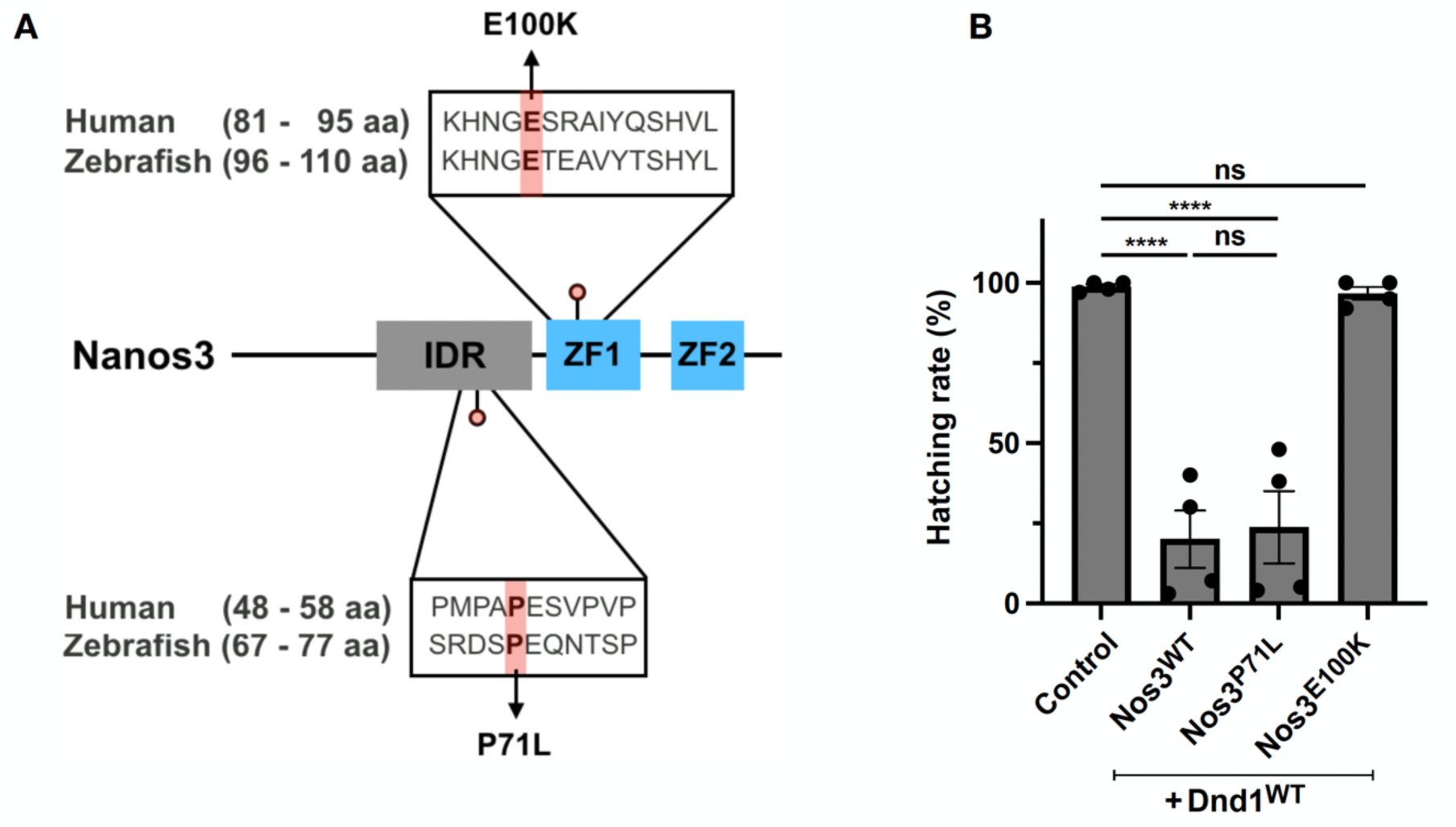
Evaluating the activity of human NANOS3 protein variants. (A) Pairwise alignment of human NANOS3 and zebrafish Nanos3 proteins, indicating the position of the amino acid exchanges in the human variants. The zebrafish Nanos3 E100K and P71L variants correspond to the human NANOS3 E85K and P52L variants, respectively. Amino acids are identified using their single-letter amino acid codes. (B) Quantification of hatching rate at 72 hpf for embryos globally co-expressing RNA encoding for the wild-type version of Dnd1 and that encoding for wild-type Nanos3 or variants (N=4, Control n = 261; Nos3^WT^ n = 222; Nos3^P71L^ n = 192; Nos3^E100K^ n = 252). ns : Not significant (P>0.05), * : P ≤ 0.05, ** : P ≤ 0.01, *** : P ≤ 0.001, **** : P ≤ 0.0001.

In contrast, co-expression of the Nanos3^E100K^ variant with Dnd1^WT^ did not affect embryonic development nor hatching rates (**Fig. 2 B**). Consistently, the E100 amino acid is located within the first conserved zinc finger domain of Nanos3 (**Fig. 2 A**), a region shown to be critical for the RNA-binding function of the protein *in vitro* and *in vivo* (Arrizabalaga and Lehmann, 1999; Curtis et al., 1997; Hashimoto et al., 2010). This is also consistent with the high CADD score of 31 of the human variant p.(Glu85Lys) (**Table 1**). Taken together, these findings indicate that Nanos3^E100K^ represents a strong loss-of-function allele, and that the corresponding human NANOS3^E85K^ protein could potentially be involved in the infertility phenotype observed in patient M2941.

### Nanos2 variant represent a partial loss-of-function allele

We next analyzed one human *NANOS2* variant employing the same assay. Accordingly, the human NANOS2 and zebrafish Nanos2 protein sequences were aligned, revealing that the variant is located within the zinc finger domain, and involves a conserved amino acid residue (**Fig. 3 A**). Using the assay presented in **Fig 1 A** we found that Nanos2^G90R^ (corresponding to human NANOS2 variant p.Gly70Arg; G70R; CADD 32) variant, result in a partial loss of function (**Fig. 3 B**). Together, these data demonstrate that co-expression of Nanos2 ^WT^ with Dnd1^WT^ in zebrafish somatic cells provides a robust readout to evaluate the activity of human NANOS2 protein variants.

**Figure 3:**
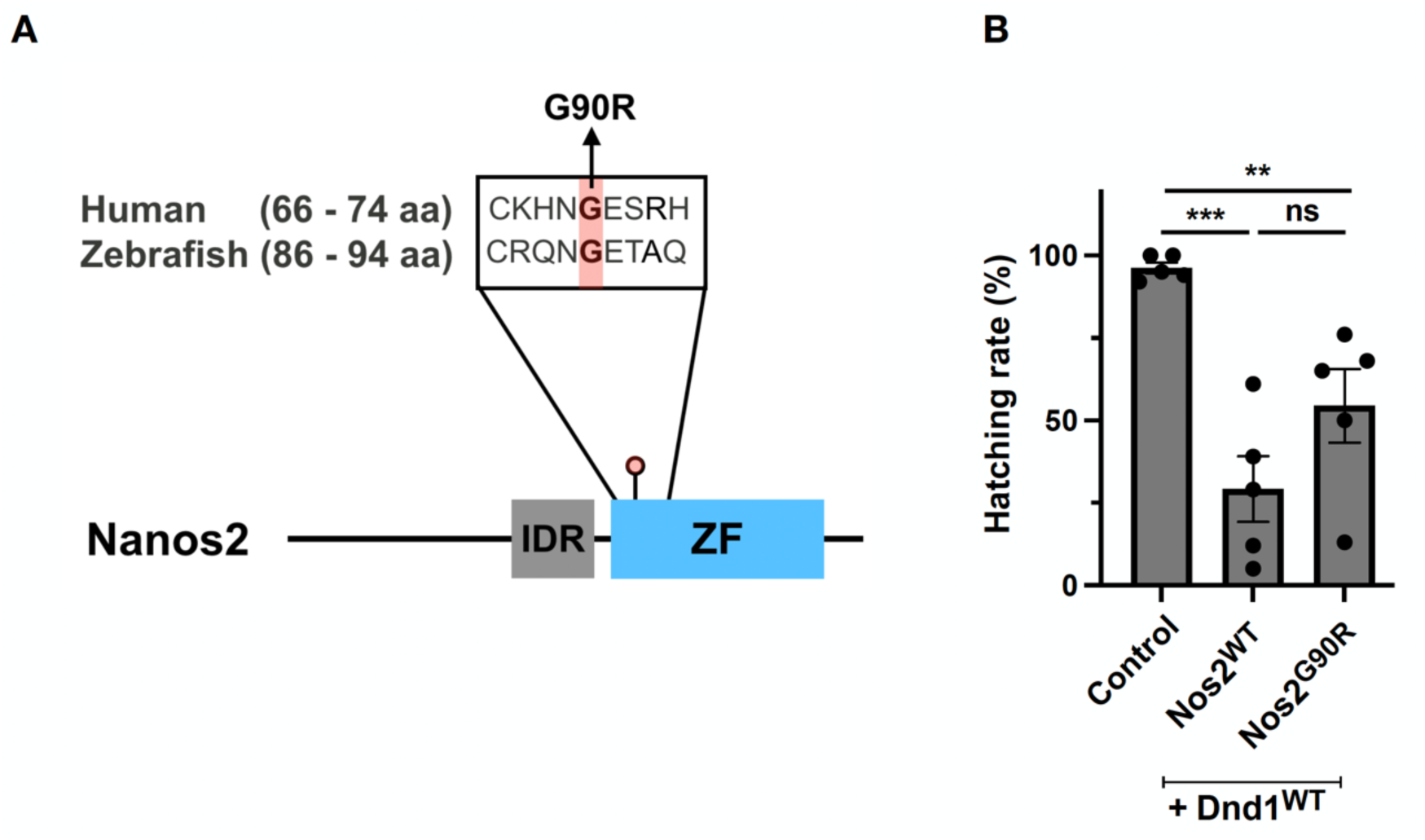
Evaluating the activity of human NANOS2 protein variants. (A) Pairwise alignment of human NANOS2 and zebrafish Nanos2 proteins, indicating the position of the amino acid exchanges in the human variants. The zebrafish Nanos2 G90R variant corresponds to human G70R variant. Amino acids are represented using their single letter amino acid codes. (B) Quantification of hatching rate at 72 hpf for embryos globally co-expressing RNA encoding for the wild-type version of Dnd1 and that encoding for the Nanos2 wild-type or variant (N=5, Control n = 286; Nos2^WT^ n = 276; Nos2^G90R^ n = 233). ns : Not significant (P>0.05), * : P ≤ 0.05, ** : P ≤ 0.01, *** : P ≤ 0.001, **** : P ≤ 0.0001.

### Validation of the Nanos3 assay using a published zebrafish Dnd1 loss-of-function variant

To check if the scheme presented above can be employed for evaluating human *DND1* variants, we co-expressed a published zebrafish Dnd1 variant - Dnd1^L329PfsTer16^ (equivalent to the human *DND1* variant p.L259PfsTer60) (Westerich et al., 2023a) with Nanos3^WT^ **(Supplementary Fig. 2A)**. Indeed, we find that embryos co-injected with *Nanos3^WT^*and *Dnd1^L329PfsTer16^* RNAs developed normally in line with the idea that this version of Dnd1 is inactive (**Supplementary Fig. 2B)**.

## Discussion

In this study, we present a rapid and robust *in vivo* assay that allows the evaluation of the activity of Nanos protein variants that were found in men with infertility. Using the proposed *in vivo* assay and the rescue experiments, we showed that Nanos2^WT^ can functionally compensate for Nanos3^WT^, consistent with previous findings by (Suzuki et al., 2007b). Using this simple yet high-throughput pipeline, human *NANOS2* and *NANOS3* variants identified in men with infertility may or may not contribute to infertility, depending on the specific variant identified. Because all identified NANOS variants were present in the heterozygous state, the mechanisms through which they may predispose to the disease remain unresolved. The variant proteins may reduce the overall functional NANOS dosage through haploinsufficiency, or act together in combination with variants at additional loci. The procedure setup presented in this work bypasses the need for biochemical tests of the protein variants for evaluating the function of the altered protein. Importantly, the approach presented here could be employed in other cases where the ectopic expression of germline-specific proteins causes defects in the development of somatic tissues.

## Materials and Methods

### Study Population

The Male Reproductive Genomics (MERGE) study currently includes the exome/genome data from 3,338 men in couples affected by infertility (Koser et al., 2026). The phenotypic spectrum encompasses normozoospermia (N=96, all semen parameters in the reference range), crypto-or azoospermia (N=577 and 2038, spermatozoa only found after centrifugation or no spermatozoa, respectively), and oligo-, astheno-and/or teratozoospermia (N=627, reduced sperm count of <39 million sperm, impaired motility and/or morphology). All men provided written informed consent, and the study was approved by the Ethics Committee of the Medical Faculty Münster and the Ärztekammer Westfalen-Lippe (2010-578-f-S, 2026-431-f-S) in accordance with the Helsinki Declaration of 1975. A targeted screen for variants in the genes NANOS1, NANOS2, and NANOS3 was performed among the members of IMIGC. Subject NOA5 belonged to a cohort of 44 patients who underwent Whole Genome Sequencing (WGS) from the Fundacio Puigvert, Barcelona, Spain, all patients signed an informed consent form and the study was approved by the local Ethical Committee of the Fundacio Puigvert, Barcelona (PI23/00425). Patient identification numbers have been double-pseudonymized, and not traceable back to the patients.

### Genetic analysis

Exome/genome sequencing from genomic DNA was performed as described previously (Koser et al., 2026). Obtained sequencing data were screened for variants in the genes *NANOS1*, *NANOS2*, and *NANOS3* according to the following filter criteria: coding, rare variants with a minor allele frequency [MAF] < 0.01 in gnomAD database v 2.1.1 (Karczewski et al., 2020), and < 0.01 in our in-house database), high impact, i.e. frameshift, stop gain, stop/start loss, splice site, and missense (CADD ≥ 20, (Kircher et al., 2014)) variants. Homozygous, potentially compound heterozygous, and heterozygous variants, which are conserved between human and zebrafish were further considered for functional analyses. The IMIGC was asked for variants considering the same filtering criteria.

Men with variants in *NANOS1*, *NANOS2*, and *NANOS3* were screened for likely pathogenic variants in valid disease genes for azoospermia and oligo-, astheno- and/or teratozoospermia (Stallmeyer et al., 2025).

### Zebrafish strains and experimental replicates

Zebrafish of the AB and AB/Tübingen long-fin (TL) genetic background were used for all co-expression experiments. For morpholino-mediated Nanos3 knockdown experiments, we used the transgenic line Tg(kop:EGFP-FTASE-UTRnanos3) (er1Tg, RRID: ZDB-ALT-070406-1, Blaser et al., 2005), in which enhanced green fluorescent protein (EGFP) is expressed at the primordial germ cell (PGC) membrane, allowing PGCs to be counted in the gonadal region. Embryos were maintained in 0.3X Danieau’s solution (17.4 mM NaCl, 0.21 mM KCl, 0.12 mM MgSO4·7H2O, and 0.18 mM Ca(NO3)2) at 28°C. All experiments were performed as independent biological replicates, and two double-blind experiments were included for all co-expression assays described in the paper.

### Cloning of DNA constructs

To generate the human NANOS protein variants for analysis in zebrafish Nanos proteins, single-nucleotide substitutions or deletions were introduced into Morpholino-resistant nanos1-globin 3′UTR, nanos2-globin 3′UTR, and nanos3-globin 3′UTR plasmids by site-directed mutagenesis, as previously described (Edelheit et al., 2009). The selected residues were based on protein sequence alignment between the two species in the relevant region, performed using Needleman–Wunsch global alignment in SnapGene version 8.2.2. We focused primarily on conserved amino acids and introduced the human variant at the corresponding position in the zebrafish protein. In each case, the substituted residue belonged to a different amino acid type than the wild-type residue.

### Microinjection into zebrafish embryos

Capped sense mRNAs were synthesized using the mMessage mMachine kit (Ambion) according to the manufacturer’s protocol. Embryos were injected at the one-cell stage with 2 nl of mRNA and/or morpholino-containing solution. For co-expression assays involving Nanos3 variants, wild-type *dnd1-globin3′UTR* mRNA together with wild-type *nanos3-*or variant *nanos3-globin 3′UTR* mRNA were injected. For co-expression assay involving Nanos2 variants, wild-type *dnd1-globin3′UTR* mRNA was co-injected with wild-type *nanos2-*or variant *nanos2-globin3′UTR* mRNA. An injection control consisting of *egfp-globin3′UTR* mRNA was co-injected to label all cells in the injected embryos and to exclude un-injected embryos from analysis. The morpholino (MO) targeting *nanos3* was injected at a final concentration of 50 µM.

### Microscopy, imaging, and statistical analysis

Imaging and counting germ cells in the gonad region were performed using a Yokogawa CSU-X1 spinning-disk microscope equipped with 20x and 63x water-immersion objectives (Zeiss) and a Hamamatsu ORCA Flash 4.0 V3 camera. The number of PGCs in the developing gonadal region was quantified in 20-22 hpf embryos, and counts were performed on one lateral side of each fish. The microscopy setup was controlled with VisiView software (Visitron, version 4.0.0.14). Brightfield images were acquired using a Leica MZ16F stereo microscope equipped with a Leica FluoCombi III™ connected to a X-Cite 120 fluorescence lamp illuminator. Imaging parameters were adjusted using Zen software, and images were captured after white balance and automatic exposure correction. Brightfield exposure times were set to 35 ms or 55 ms. Statistical analyses were carried out in GraphPad Prism (version 9.4.0). Each experiment was performed as independent replicates, including two double-blind experiments, and significance was assessed using Ordinary one-way ANOVA (Tukey’s multiple comparisons test), with P < 0.05 considered statistically significant. ns : Not significant (P>0.05), * : P ≤ 0.05, ** : P ≤ 0.01, *** : P ≤ 0.001, **** : P ≤ 0.0001.

## Data Availability

All data produced in the present study are available upon reasonable request to the authors

## Trial registration number

N/A.

---

**Supplementary Figure 1.**
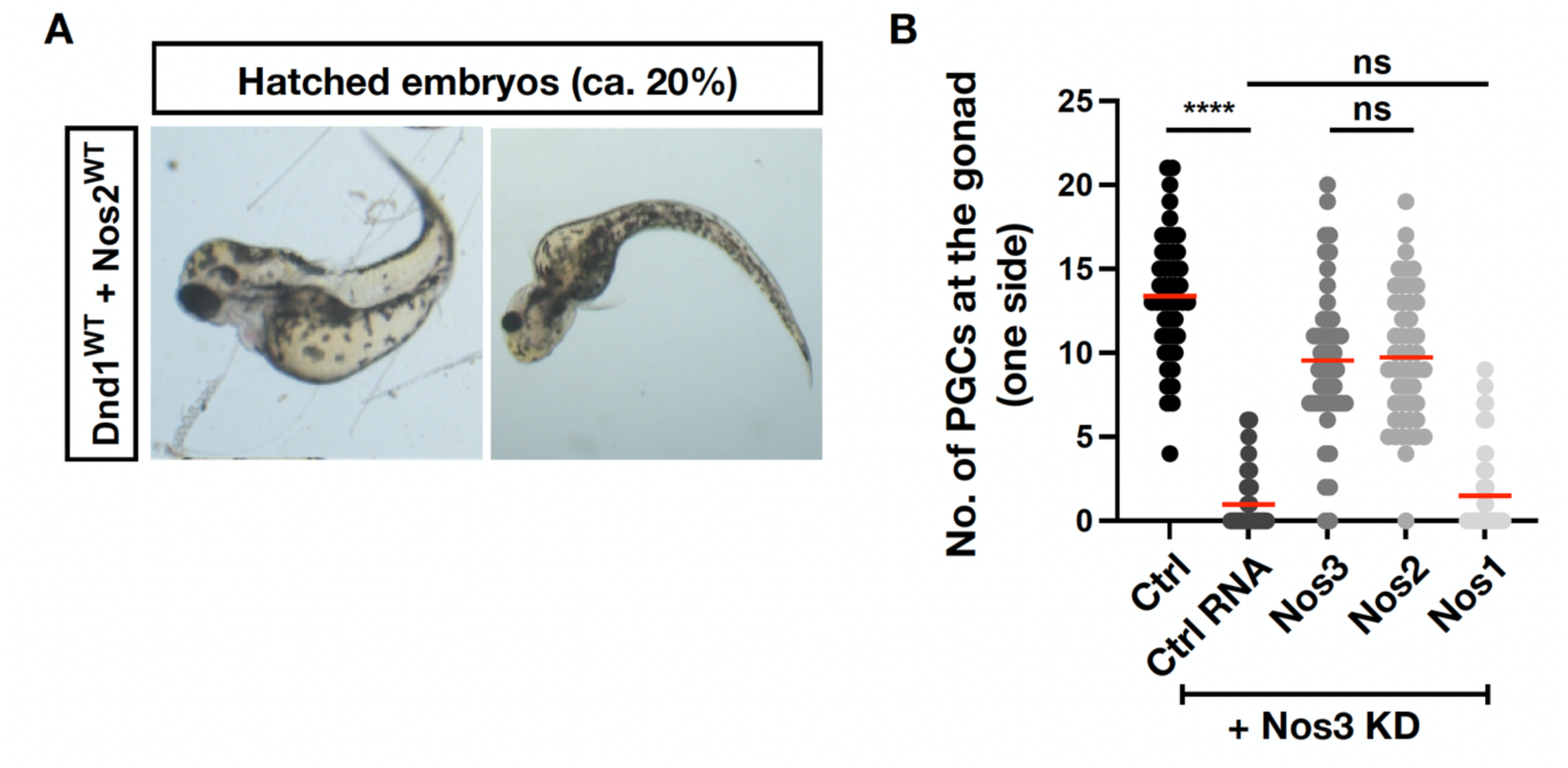
(A) Examples of hatched 72 hpf embryos following co-injection of Nanos2^WT^ and Dnd1^WT^ encoding RNAs. (B) Reversal of the Nanos3-morpholino (MO) induced knockdown of Nanos3 by injection of *control*, *nanos3, nanos2, nanos1* mRNAs as assayed by the number of PGCs located at the region where the gonad develops. As a control - control MO was injected along with *control* mRNA. The number of PGCs at the gonad was determined at 20-22 hpf (N=3, MO-control n = 48; Control RNA n = 48; Nos3^WT^ n = 48; Nos2^WT^ n = 49; Nos1^WT^ n = 49).

**Supplementary Figure 2.**
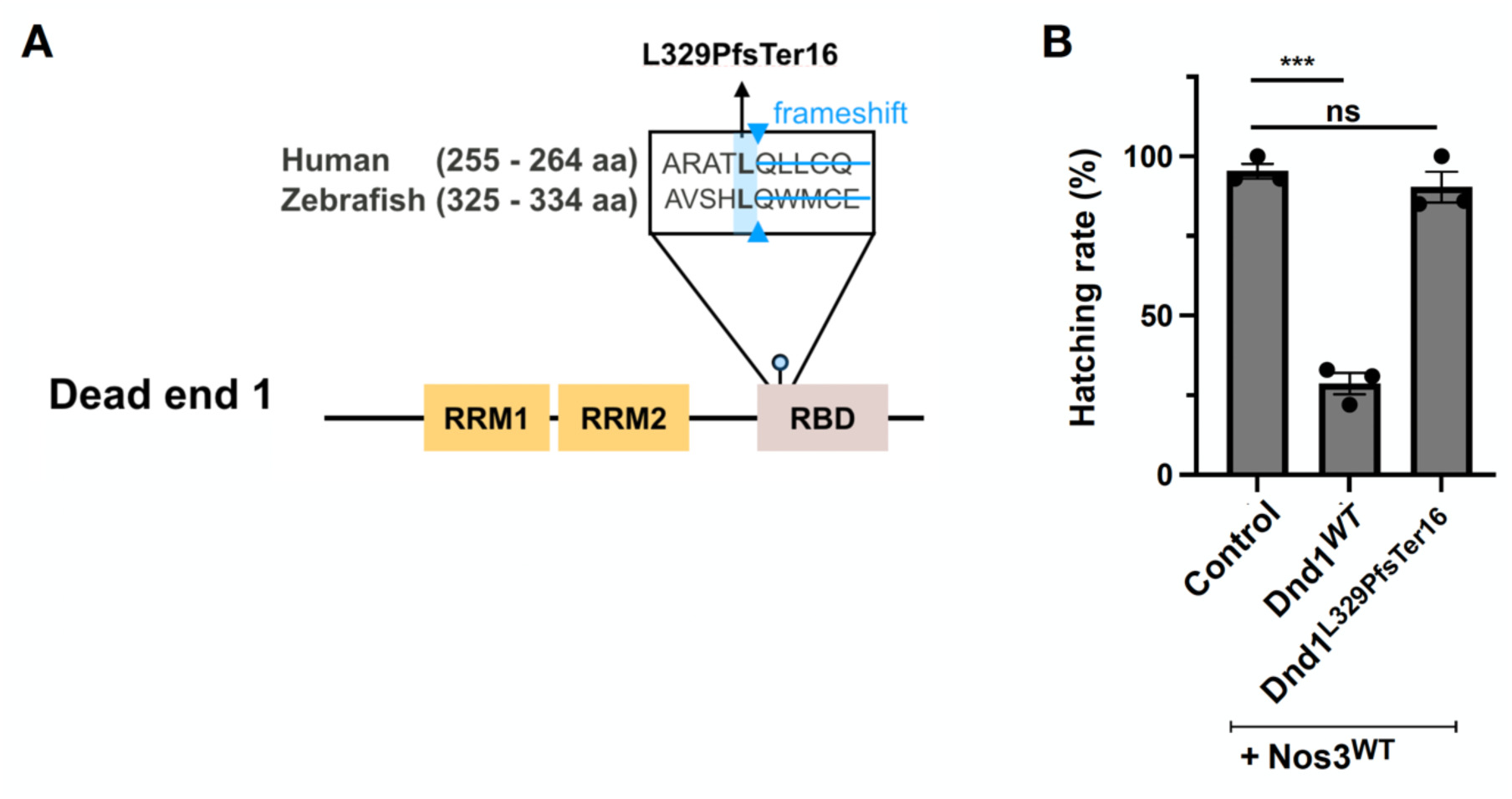
(A) Schematic of the pairwise alignment of human DND1 and zebrafish Dnd1 proteins. The zebrafish L329PfsTer16 corresponds to human L259PfsTer60. Amino acids are represented using their single letter amino acid codes. (B) Quantification of hatching rate at 72 hpf for embryos globally co-expressing RNA encoding for the wild-type version of Nanos3 and that encoding for the Dnd1 wild-type or variant (N=3, Control n =125; Dnd1^WT^ n = 120; Dnd1^L329PfsTer16^ n = 118).

